# Lipoprotein(a) and large artery atherosclerosis: Results from the BIOSIGNAL Study

**DOI:** 10.1101/2024.08.14.24312026

**Authors:** Felix Gross, Valerie Schütz, Laura Westphal, Corinne Inauen, Thomas Pokorny, Susanne Wegener, Andreas R. Luft, Katharina Spanaus, Arnold von Eckardstein, Pierre-Jean Touboul, Markus Arnold, Mira Katan

## Abstract

**Background:** Lipoprotein(a) (Lp(a)) is a causal risk-factor for atherosclerotic cardiovascular disease including acute ischemic stroke (AIS). The underlying pathomechanisms mediating this risk are less well understood, especially in AIS caused by large artery atherosclerosis (LAA). In this observational cohort study we evaluated the association of Lp(a) with markers of LAA, namely carotid intima media thickness (cIMT) and the presence of extra- or intracranial vessel narrowing plaques.

**Methods:** Among participants of the BIOSIGNAL cohort study we determined Lp(a) levels within 24h after symptom onset in 1161 AIS patients from the single center of Zurich. cIMT was determined using a semi-automated computerized edge tracking software, internal carotid artery (ICA) stenosis was graded according to the North American Symptomatic Carotid Endarterectomy Trial (NASCET) criteria, intracranial ultrasound was performed by transcranial color-coded duplex (TCCD).

**Results:** In 1161 participants higher Lp(a) levels were not associated with an increased cIMT in univariable or multivariable regression models containing known cardiovascular risk-factors. Higher Lp(a) levels were not associated with the presence of neither extracranial high-grade ICA-stenosis nor significant intracranial stenosis assessed by neurovascular ultrasound.

**Conclusion:** In AIS patients higher Lp(a) levels were not associated with clinical markers of arteriosclerotic burden despite its association with LAA-stroke etiology and an increased risk for stroke recurrence.

**Registration:** Registration-URL: http://www.clinicaltrials.gov; Unique identifier: NCT-02274727

## Introduction

Lp(a) resembles to low-densitiy lipoprotein (LDL) by the presence of apolipoprotein B-100 and enrichment with cholesteryl esters. It differs from LDL by the additional presence of apolipoprotein (a) (apo(a)) which shares structural similarities with plasminogen. Lp(a) has been established as a genetically determined, independent, and likely causal risk-factor for atherosclerotic cardiovascular disease (1). Currently, several RNA-based therapeutics targeting Lp(a) levels are undergoing clinical development (2). Despite the well-established association between Lp(a) and atherosclerotic cardiovascular disease (ASCVD), the relationship between Lp(a) and ischemic stroke remains inadequately understood, particularly in the context of secondary prevention. While incident ischemic stroke has been investigated as an outcome measure in cohorts with cardiovascular risk as well as population-based studies (3), limited research has explored the association of Lp(a) with stroke recurrence following an index event (4). Recent findings have demonstrated that Lp(a) levels ≥100 nmol/l are associated with an increased risk for recurrent events among patients who are either <60 years or present with evident carotid arteriosclerotic disease (5), prompting further investigation into the potential role of Lp(a) lowering in secondary stroke prevention.

However, the underlying pathophysiological mechanisms mediating the association of elevated Lp(a) levels with an increased stroke risk remain a subject of debate. Given the structural similarities between Lp(a) and LDL as well as apo(a) with plasminogen, both pro-atherosclerotic and prothrombotic/antifibrinolytic properties of Lp(a) have been discussed as potential underlying mechanisms in vivo (6). In addition, the enrichment with oxidized phospholipids in Lp(a) suggested a proinflammatory role. A better comprehension of the pathophysiological mechanisms operative in distinct cohorts is crucial to identify target populations in forthcoming medical interventions. This is particularly significant as the impact on pro-arteriosclerotic versus pro-thrombotic/antifibrinolytic properties is likely to manifest over different periods of time.

The aim of this study is to determine the association between elevated Lp(a) levels and markers of arteriopathy, specifically carotid intima media thickness (cIMT), and the extent of extracranial carotid artery stenosis and intracranial artery stenosis. To achieve this, we analyzed a well-described cohort of patients with acute ischemic stroke (AIS), where Lp(a) levels were measured at a standardized time point following the event. Extra- and intracranial artery stenosis was evaluated by vascular ultrasound and cIMT was estimated using a semi-automated software solution.

## Methods

### Data Availability

The de-identified data supporting the findings of this study are available from the corresponding author on reasonable request.

### Study Protocol Approvals, Registrations and Patient Consents

The BIOSIGNAL (Biomarker Signature of Stroke Aetiology) study (ClinicalTrial.gov NCT02274727) was approved by local ethics committees and conducted according to the principles expressed in the Declaration of Helsinki. The protocol is in adherent to the STROBE observational cohort guidelines.(7) All patients or their welfare guardians provided written informed consent for the collection of blood, collection of data and subsequent analyses.

### Study Design and cohort description

The BIOSIGNAL study is a prospective, observational, multicenter, inception study to evaluate selected blood biomarkers in patients with acute ischemic stroke measured within 24 hours from symptom onset which has been described in detail elsewhere (5). Briefly, from October 2014, through October 2017, a total of 1759 patients who were admitted with ischemic stroke were enrolled. Ischemic stroke was defined according to the World Health Organization criteria as an acute focal neurological deficit lasting longer than 24 hours. If symptoms did not persist beyond 24 hours, cranial imaging confirmed a new ischemic infarct. Patients with hemorrhagic stroke, TIA lacking tissue-based confirmation of ischemia or patients discharged from the hospital with a diagnosis different from ischemic stroke (i.e. stroke mimics) were excluded. For this single-center analysis we included a subcohort containing 1161 patients enrolled at the University Hospital Zurich.

### Standardized Patient Work-up

All participants received CT or MRI and routine laboratory tests on admission. Demographic variables, vital signs and vascular risk factors were collected on admission. Hypertension was defined as blood pressure above 140/90 mmHg repeatedly or current medical treatment with antihypertensive agents. Dyslipidemia was defined as LDL levels above 2,6 mmol/L or current medical treatment with lipid-lowering drugs. Active smoking status was defined as reported current smoking or ceased smoking within the previous 2 years. A positive family history was defined as a cardiovascular event (stroke or myocardial infarction) in a 1^st^ grade relative ≤ 65 years. Participants received standard of care etiological workup including 12 lead electrocardiography (ECG) on admission, extra- and intracranial vascular ultrasound, prolonged ECG monitoring and transthoracic or transesophageal echocardiography. Stroke etiology was determined according to the TOAST (trial of Org 10172 in Acute Stroke Treatment) classification (8) as well as the SSS-TOAST System (9).

### Ultrasound Assessment of Atherosclerotic Burden

Extra- and intracranial ultrasound was performed during the index hospitalization in a standardized manner (10) by a board trained vascular neurologist. Grading of extracranial ICA-stenosis was performed according to the North American Symptomatic Carotid Endarterectomy Trial (NASCET) criteria (11). Ultrasound examination of intracranial arteries was performed by transcranial color-coded duplex (TCCD) via transtemporal (middle cerebral artery (MCA), anterior cerebral artery (ACA), posterior cerebral artery (PCA)) and suboccipital window (intracranial segment of vertebral arteries (V4), basilar artery (BA)) with use of ultrasound contrast agent (SonoVue) where sufficient visualization could not be achieved otherwise. Grading of intracranial stenosis (</>50%) was done according to the 2008 Consensus recommendations (12). cIMT was determined using a semi-automated computerized edge tracking software M’Ath (Intelligence in Medical Technologies, Inc., Paris, France) (13). Measurements were obtained outside the areas of plaque on the far wall at the distal common carotid artery or the bulb as recommended (14) by high-resolution B-mode ultrasound with lateral probe incidence (Siemens Acuson, 12.0 MHz Probe). cIMT was measured on 10mm in length, quality index had to be ≥ 0.3, meaning that at least 30% of measurements were available for IMT averaging. The arithmetic mean of left and right sided measurement was calculated, and this metric was used for the analysis. Whenever only one side met the quality criteria, we used the single-sided measurement value for cIMT.

### Biomarker measurement

Blood was drawn within 24 hours of symptom onset during the first routine blood sampling in EDTA containing plastic tubes. Samples were immediately centrifuged at 3000g at 4°C for 20 minutes, aliquoted, and frozen at -80°C until the time of analysis. Lp(a) levels were assessed in batches of thawed plasma with the Roche Tina-quant Lipoprotein(a) assay on the Cobas c702 platform (Roche Diagnostics, Mannheim, Germany). Plasma levels are expressed in nanomole per liter (nmol/l). The manufacturer defined the lower limits of quantification (LLoQ) and detection (LoD) as 20 and 7 nmol/l, respectively. For measurements below LLoQ (49%) we used the apparent concentrations recorded by the assay as suggested for epidemiological studies (15). Apparent concentrations below LoD (21.5%) were assigned a value of LoD divided by the square root of two as recommended previously (16). The intra assay and inter assay coefficient of variation, defined as the ratio of the SD to the mean, was < 5%, respectively. To dichotomize Lp(a) we used a cut-off of 100 nmol/L (1,5).

### Statistical Analysis

Discrete variables are summarized as counts (percentages) while continuous variables are shown as medians (interquartile ranges [IQR]). Common logarithmic transformation (base 10) was performed to transform to normality for skewed distributions. For two-group comparisons, Fisher’s exact test was employed for binary outcomes, while Mann–Whitney U test was used for continuous outcomes. For multigroup comparison the Kruskal-Wallis test was used with appropriate post-hoc testing. To investigate the association of Lp(a) with cIMT as well as the presence of extra- and/or intracranial stenosis linear and binary logistic regression models were constructed respectively to calculate regression coefficients and odds ratios (OR) with 95% confidence intervals (95% CI). Multivariate models were built using demographic variables (age, sex) and established vascular risk factors (arterial hypertension, diabetes mellitus, dyslipidemia, active smoking, history of stroke and body mass index (BMI)) as well as LDL-C levels. P-values < 0.05 were considered statistically significant. Data analysis was performed using STATA version 17 (StataCorp LLC, College Station, Texas).

## Results

### Baseline Cohort Characteristics

A total of 1166 patients with an AIS were consecutively enrolled in the study. Median age was 74 (IQR 62 – 83) years. Blood samples collected within 24 hours after symptom onset were available in 1161 patients (99.6%). Median Lp(a) levels were 16 nmol/l (IQR 7-56), 192 (16.5 %) participants had elevated Lp(a) levels (≥ 100nmol/l). 1067 (91.9%) patients received standardized neurovascular ultrasound examinations to assess extra- and intracranial arteriosclerotic burden. cIMT measurements were obtained in 964 patients on the left side and 951 patients on the right side. 888 measurements met the quality criteria for the left carotid artery and 830 for the right side respectively, resulting in an overall 955 patients (82.3%) with cIMT measured with sufficient quality on at least one side.

**Table 1:**
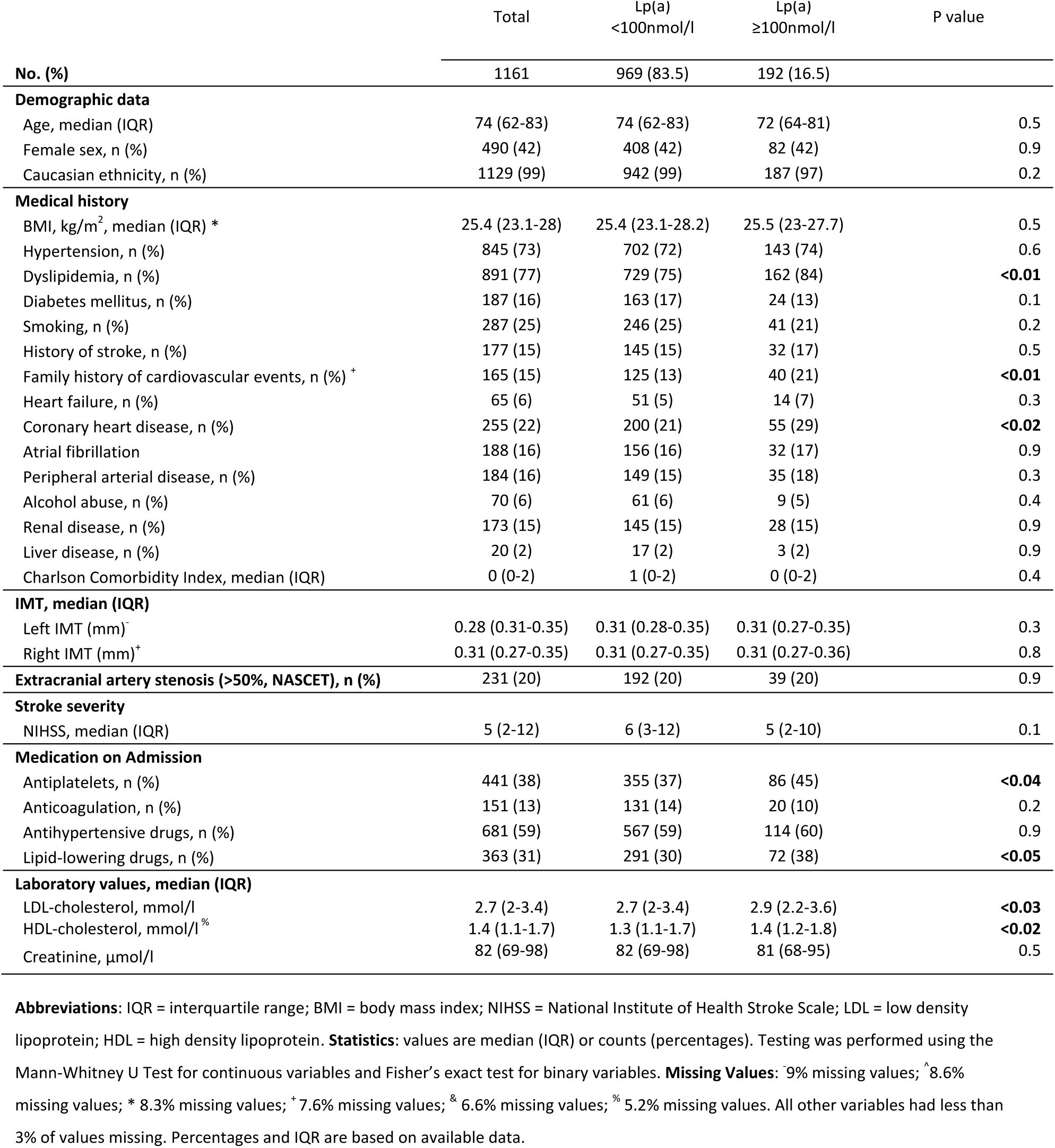
Baseline characteristics stratified by Lipoprotein (a) *≥*100nmol/l / <100nmol/l.

### Association of Lipoprotein(a) with cIMT

In linear regression analysis log_10_Lp(a) values were not associated with cIMT of the left or right carotid arteries or when both sides were combined (Reg. Coeff. -0.005, 95% CI -0.02-0.01, p=0.63 for the left side, Reg. Coeff. -0.005, 95% CI -0.02-0.15, p=0.6 for the right side, Reg. Coeff. -0.007, 95% CI -0.02-0.01, p=0.4 for both sides in univariate analysis).

Multivariate analysis did reveal a significant association between combined cIMT and age, hypertension and LDL-C levels (p=0.00, p=0.01 and p=0.01 respectively in the full model.

Figure 1 shows (cIMT) in mm stratified by age group and dichotomized Lp(a) (<100nmol/l vs. >100nmol/l)

**Figure 1:**
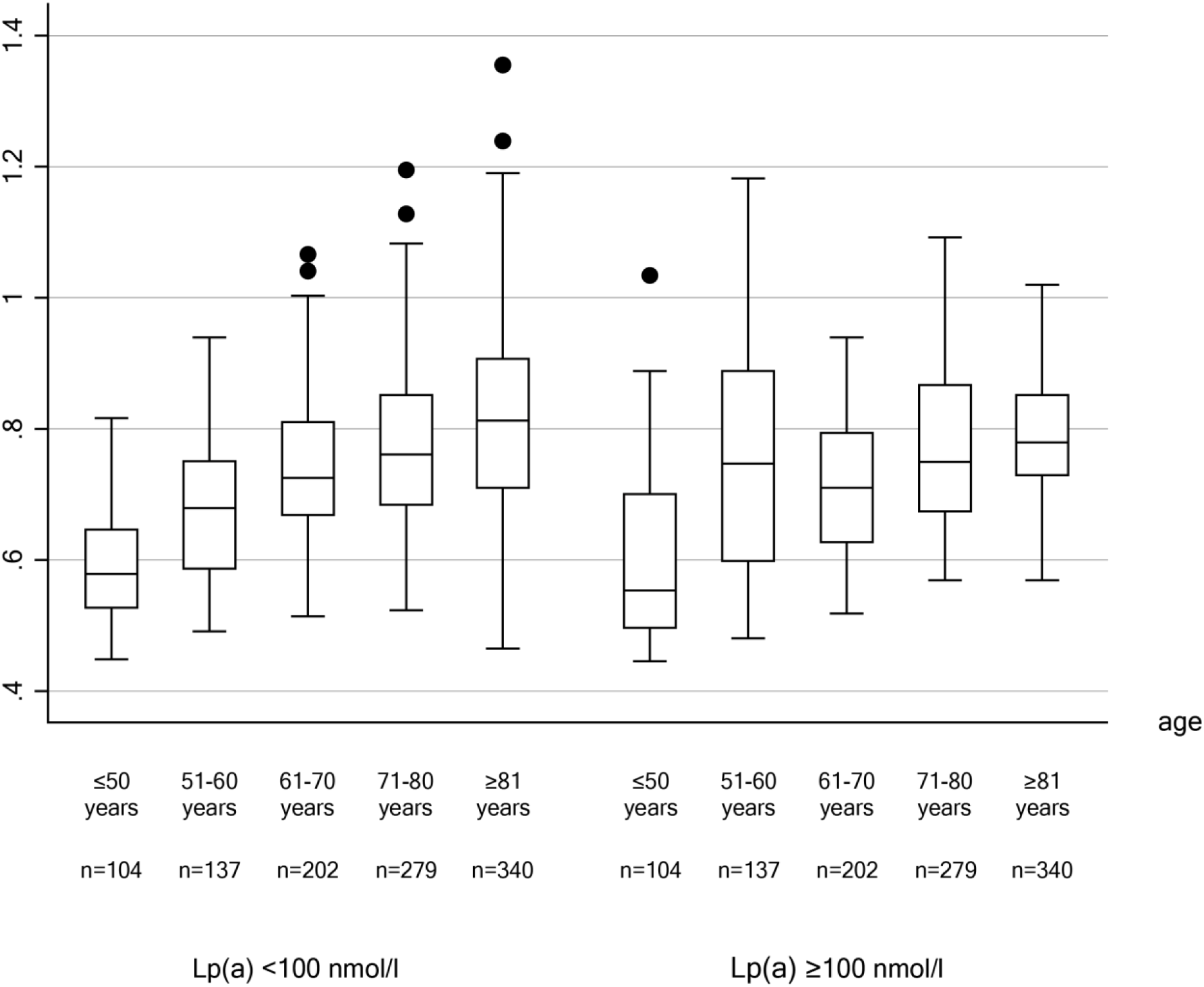
Carotid Intima media thickness (cIMT) in mm stratified by age group and dichotomized Lp(a) Boxplot of carotid intima media thickness (cIMT) in milimeters in participants with Lp(a) levels below and above 100nmol/l stratified by agegroups with median and 25^th^ and 75^th^ percentile. The whiskers extend to the last data point beyond 1.5 * the interquartile Range, circles represent outliers.

### Association of Lipoprotein(a) with extracranial vessel stenosis

According to the SSS-TOAST criteria 176 (15.2%) of index events were classified as large-artery atherosclerotic (LAA) stroke-subtype. Lp(a) values were numerically higher in patients with LAA stroke etiology compared to other subtypes, but this association was not statistically significant (OR 1.33, 95%CI 1.02-2.12, p=0.053 for log transformed Lp(a)) in the analyzed cohort. In 231 patients at least one ≥50% ICA-stenosis according to NASCET criteria was found (left side n=158, right side n=144). In logistic regression analysis log_10_Lp(a) values were not associated with the observation of ≥50% left ICA-stenosis (OR 1,06, 95%CI 0.78-1.44, p=0.72), ≥50% right ICA-stenosis (OR 1.24, 95%CI 0.91-1.71, p=0.17) or when both sides were combined (OR 1.11, 95% CI 0.86-1.43, p=0.42).

252 (21.7%) patients had ≥50% stenosis of at least one intracranial artery. Lp(a) levels were not associated with the occurrence of intracranial stenosis (OR 0.83, 95%CI 0.66-1.05, p=0.116). Results did not change after exclusion of patients with significant extracranial stenosis to prevent misclassification. In linear regression analysis log_10_Lp(a) values were not associated with extra- and intracranial stenosis combined.

Figure 2 represents Lp(a) in nmol/l stratified by ultrasound confirmed extra- and/or intracranial arterial stenosis.

**Figure 2:**
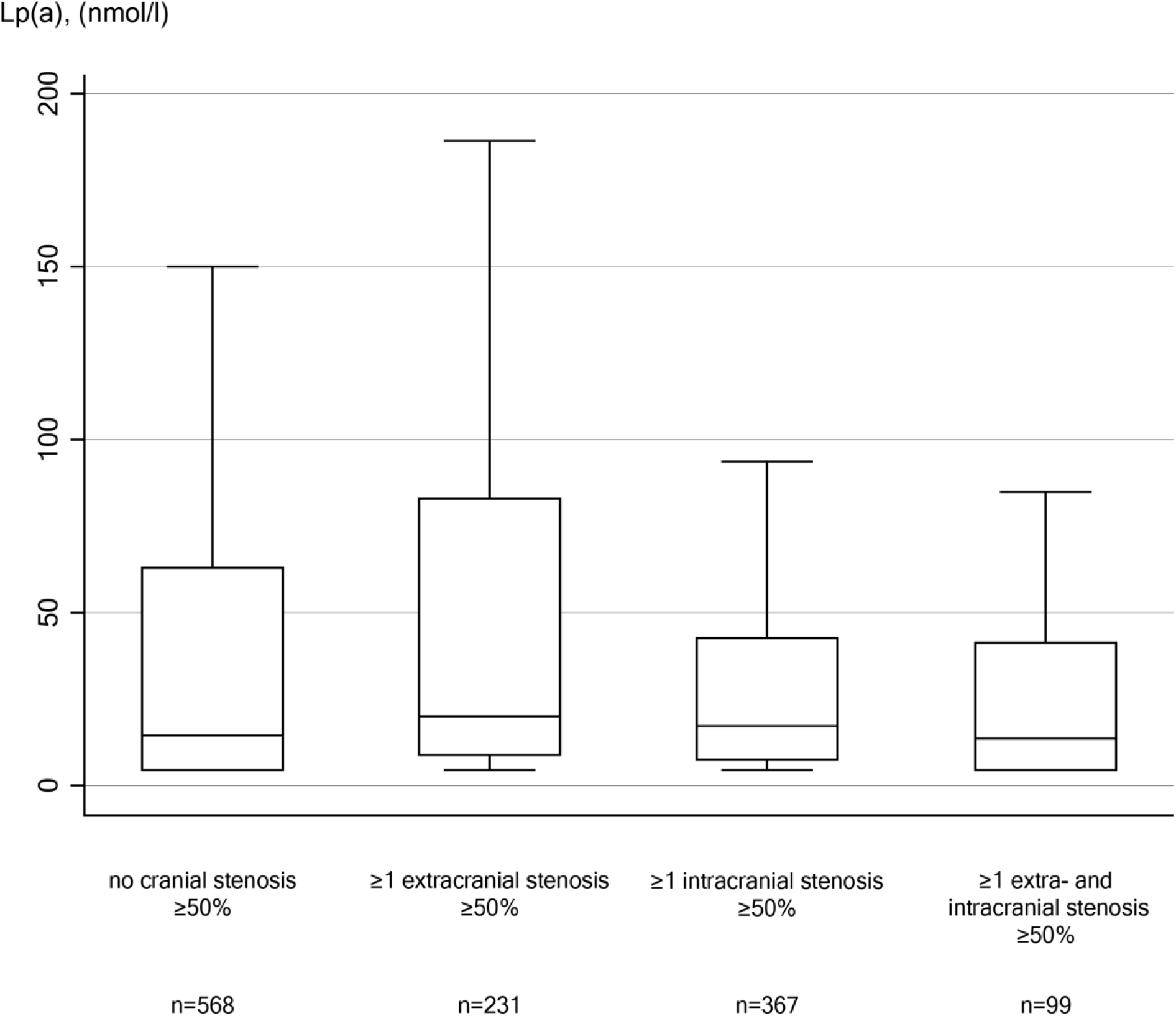
Lp(a) in nmol/l stratified by ultrasound confirmed extra- and/or intracranial arterial stenosis. Box plot of Lipoprotein(a) levels (Lp(a)) in nmol/l by number of extra- and or intracranial arterial stenosis with median and 25^th^ and 75^th^ percentile. The whiskers extend to the last data point beyond 1.5 * the interquartile Range.

## Discussion

We previously reported that Lp(a) levels (>100nmol/l) measured within 24h after ischemic stroke were associated with an increased risk of stroke recurrence, especially in younger stroke patients and if large-artery arteriosclerosis was present (5). In the present analysis, we investigated the association of Lp(a) levels with sonographic markers of cerebrovascular arteriopathy, namely cIMT and the presence of intra- or extracranial artery stenosis.

Despite the robust association of Lp(a) levels with LAA stroke etiology, which has also been confirmed in a recent meta-analysis (17), we could not demonstrate any association of elevated Lp(a) with evident atherosclerotic burden on ultrasound imaging. These results may indicate that the effect of Lp(a) on stroke recurrence may not be solely attributable to the pro-atherogenic properties of Lp(a).

The exact pathomechanisms mediating the association of Lp(a) with an increased risk for cardiovascular events remain a topic of debate. Similar to LDL, the apolipoprotein B containing and cholesteryl ester rich Lp(a) could initiate the formation of atherosclerotic lesions after entering the arterial wall (18). Further the presence of oxidized phospholipids on apolipoprotein(a) mediates inflammation and thus seems to promote atherosclerosis (19–21).

Because of its structural homology to plasminogen apolipoprotein(a) has also been suggested to promote thrombosis (6,22). Considering intima-media thickness as a marker of arteriopathy other studies have also failed to demonstrate a clear link to elevated levels of Lp(a)(23)(24). Most recently, in a longitudinal analysis of the Cardiovascular Risk in Young Finns Study (YFS), the finding of elevated Lp(a) levels in adolescents was not associated with increased carotid artery thickness but with clinical manifest cardiovascular disease in adulthood. (25).

Considering clinically manifest atherosclerosis the existing data is conflicting. In the prospective Bruneck Study including 826 individuals, elevated Lp(a) was confirmed as a risk factor for development of incident atherogenesis synergistically with elevated LDL levels (26,27). However, in this study elevated Lp(a) was also associated with advanced stages of atherosclerosis, i.e. progression of atherosclerotic plaque into clinically relevant stenosis during 5-year follow-up. High grade stenosis often develops though plaque rupture and thrombosis rather than sole atherogenesis, and the association between elevated Lp(a) and stenosis progression was independent of LDL levels but only seen for small apo(a) isoforms. These smaller isoforms are known to have a higher antifibrinolytic capacity than larger isoforms. Thus, these results suggest that prothrombotic/antifibrinolytic properties of Lp(a) may be involved. In line with this hypothesis a 2008 study including 876 patients from an atherosclerosis prevention clinic found elevated Lp(a) associated with high grade stenosis and occlusion, but no association was found with carotid plaque area (28). Further, in the PARISK-study including 182 patients with symptomatic carotid artery stenosis it was shown that increased Lp(a) was also associated with imaging markers of plaque vulnerability, namely intraplaque hemorrhage, lipid-rich necrotic core and thin-or-ruptured fibrous cap. (29) Coincidentally, in an analysis of the AIM-HIGH study, among 214 patients with established vascular disease elevated Lp(a) was associated with high-risk plaque features on MRI (30). While the association with atherosclerotic burden in coronary arteries and severeness of aortic valve stenosis is well documented, evidence of a possible association with extra- and intracranial atherosclerosis in stroke patients is scarce. In a Korean study including 1012 consecutive stroke patients the highest quartile of Lp(a) was associated with combined extra- and intracranial stenosis (OR 4.98, 95% CI 1.92–12.91) (31).

In summary, together with the results we have documented in this large high-risk cohort of AIS patients the current available clinical data suggest that Lp(a) may not mediate its effects on stroke recurrence by initiation of atherosclerosis alone, but rather on an interplay of atherosclerotic, pro-thrombotic and inflammatory effects.

Finally, we acknowledge strengths and limitations of the presented work. Our cohort being formed by the inclusion criteria of an ischemic cerebrovascular event, we reckon it a highly specific cohort differing substantially from other cohorts in the field of Lp(a) research that generally studied coronary heart disease and aortic valve stenosis. Using a validated algorithm-assisted system for IMT-measurement allowed us to have less interrater-dependent bias and robust data for our calculations. Additionally, having measured Lp(a) values standardized within 24 hours of the index hospitalization is valuable as Lp(a) levels rise during severe illness (32). We used a second-generation Lp(a) assay for measurements, rendering results independent of highly variable apo(a) size (33). Due to either missing data or inadequate image quality for the semi-automated software, cIMT measurements were unavailable for some patients, affecting statistical power. Studies utilizing alternative modalities such as computed tomography angiography or magnetic resonance angiography may offer advantages in mitigating these limitations.

## Conclusion

Previous research has demonstrated a significant correlation between elevated Lp(a) levels and LAA etiology, as well as recurrent stroke. However, our investigation did not uncover evidence supporting a link between higher Lp(a) levels and the extent of extra- and/or intracranial atherosclerotic burden, as measured through ultrasound methods in these patients. It is likely that other mechanisms – such as prothrombotic and antifbrinolytic properties are more influential. Thus, the identification of stroke patients who could benefit from Lp(a) screening and potential intervention by novel drugs should not rely solely on the extent of stenosis. In the interim of further research to understand the pathomechanisms of Lp(a) on stroke incidence, the management of other recognized and modifiable cardiovascular risk factors remains crucial in the day-to-day care and counseling of patients.

## Data Availability

The deidentified data supporting the findings of this study are available from the corresponding author on reasonable request.

